# Alpha globin gene copy number and incident ischemic stroke risk among Black Americans

**DOI:** 10.1101/2023.03.15.23286908

**Authors:** A. Parker Ruhl, Neal Jeffries, Yu Yang, Steven D. Brooks, Rakhi P. Naik, Lydia H. Pecker, Bryan T. Mott, Cheryl A. Winkler, Nicole D. Armstrong, Neil A. Zakai, Orlando M. Gutierrez, Suzanne E. Judd, Virginia J. Howard, George Howard, Marguerite R. Irvin, Mary Cushman, Hans C. Ackerman

## Abstract

**Introduction:** People with African ancestry have greater stroke risk and greater heritability of stroke risk than people of other ancestries. Given the importance of nitric oxide (NO) in stroke, and recent evidence that alpha globin restricts nitric oxide release from vascular endothelial cells, we hypothesized that alpha globin gene (*HBA)* deletion would be associated with reduced risk of incident ischemic stroke.

**Methods:** We evaluated 8,947 participants self-reporting African ancestry in the national, prospective Reasons for Geographic And Racial Differences in Stroke (REGARDS) cohort. Incident ischemic stroke was defined as non-hemorrhagic stroke with focal neurological deficit lasting ≥ 24 hours confirmed by the medical record or focal or non-focal neurological deficit with positive imaging confirmed with medical records. Genomic DNA was analyzed using droplet digital PCR to determine *HBA* copy number. Multivariable Cox proportional hazards regression was used to estimate the hazard ratio (HR) of *HBA* copy number on time to first ischemic stroke.

**Results:** Four-hundred seventy-nine (5.3%) participants had an incident ischemic stroke over a median (IQR) of 11.0 (5.7, 14.0) years’ follow-up. *HBA* copy number ranged from 2 to 6: 368 (4%) -α/-α, 2,480 (28%) -α/αα, 6,014 (67%) αα/αα, 83 (1%) ααα/αα and 2 (<1%) ααα/ααα. The adjusted HR of ischemic stroke with *HBA* copy number was 1.04; 95%CI 0.89, 1.21; p = 0.66.

**Conclusions:** Although a reduction in *HBA* copy number is expected to increase endothelial nitric oxide signaling in the human vascular endothelium, *HBA* copy number was not associated with incident ischemic stroke in this large cohort of Black Americans.

## INTRODUCTION

People with African ancestry have greater stroke risk and greater heritability of stroke risk than people of other ancestries.^1,2^ While some risk is explained by genetic variants shared between European and African populations, less is known about African-specific variants.^3^ Alpha globin (*HBA*), which regulates endothelial nitric oxide (NO) signaling in resistance arteries, varies in gene copy number among people of African and Asian descent.^4,5^ Genetic deletion of *HBA* is associated with protection from kidney disease, possibly through increased vascular NO signaling.^6^

NO plays an important role as a signaling molecule which regulates cerebrovascular blood flow both at rest and during injury.^7^ The regulation of cerebrovascular blood flow via NO occurs via two main mechanisms: 1) autoregulation to keep a consistent blood flow and 2) neurovascular coupling in which increased neuronal activity is influenced by the local regulation of cerebral blood flow. When there is reduced cerebral blood flow, for example during either an ischemic stroke or a hemorrhagic stroke, depletion of NO may exacerbate neuronal injury. However, the role of NO in the cerebral vasculature is complex and an overabundance of NO leading to significant vasodilation and lower cerebral perfusion pressure could potentially contribute to cerebral injury during stroke event. In addition, NO may impact the risk of developing vasospasm in the setting of cerebral hemorrhage.^8^ Given the importance of NO in stroke,^7,9^ and evidence that the alpha subunit of hemoglobin restricts nitric oxide release from vascular endothelial cells, we examined the association between *HBA* copy number and incident ischemic stroke in the Reasons for Geographic And Racial Differences in Stroke (REGARDS) cohort.^10^ We hypothesized that lower *HBA* copy number would be associated with lower risk of incident ischemic stroke.

## METHODS

### Study Design

REGARDS is a longitudinal cohort study designed to determine the reasons for racial disparities in stroke and cognitive decline in Black and White Americans aged ≥ 45 years.^10^ REGARDS enrolled 30,239 participants from the 48 continental United States from 2003 to 2007. All self-reported Black participants consenting to genetic research were included in this study (Figure 1). All participants provided oral and written informed consent. The REGARDS study was approved by the Institutional Review Boards of participating centers. This study followed the Strengthening the Reporting of Observational Studies in Epidemiology (STROBE) reporting guideline. The analytic plan was prespecified and approved by the REGARDS Publications Committee. Among 9,999 Black REGARDS participants with *HBA* and *HBB* genotyping available, 8,947 participants had no self-reported history of stroke at baseline and complete follow-up data (Figure 1).

**Figure 1.**
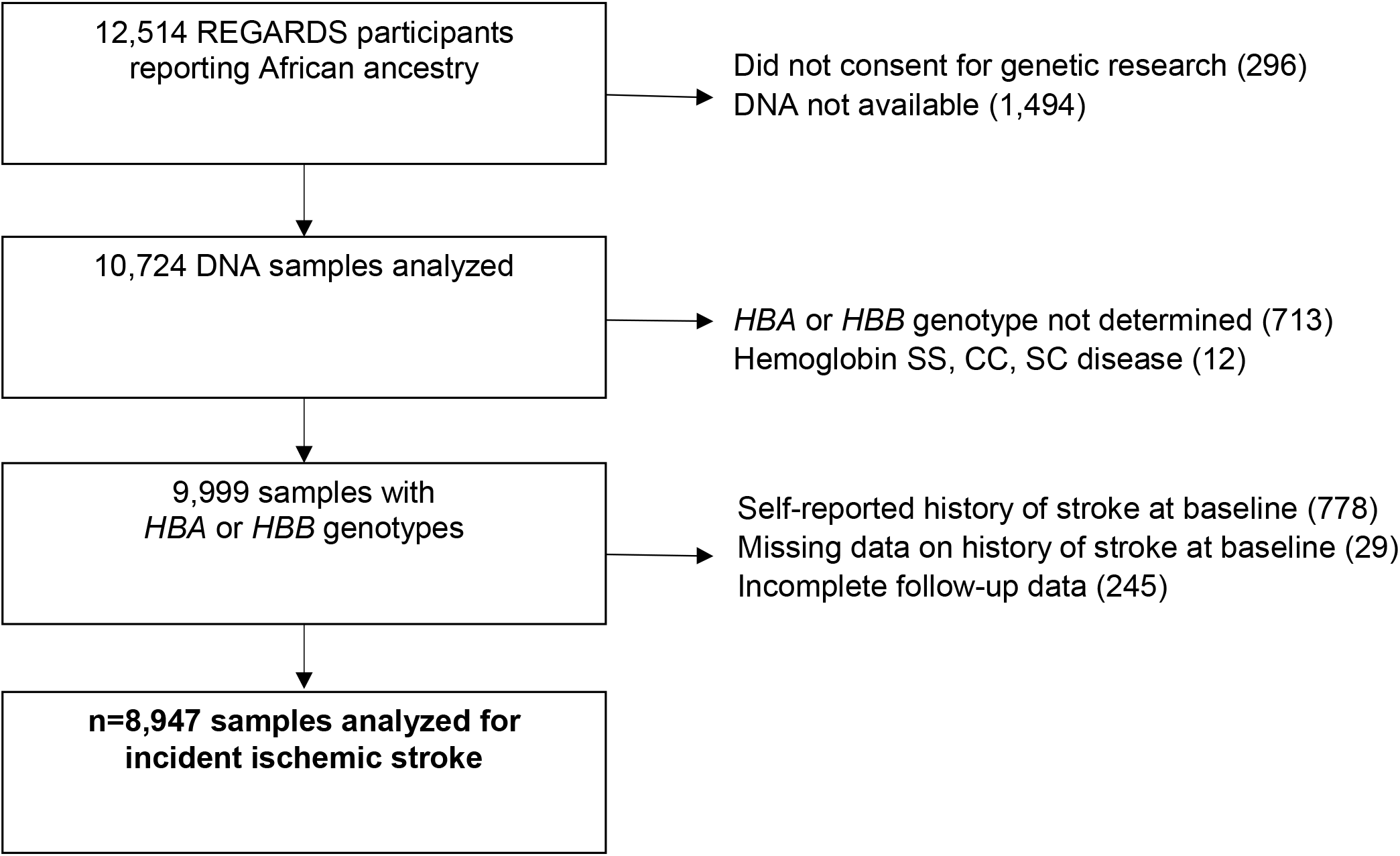
Incident ischemic stroke and *HBA* copy number cohort study flow diagram REGARDS = Reasons for Geographic and Racial Differences in Stroke (REGARDS) longitudinal cohort study; *HBA* = alpha globin gene; *HBB* = beta globin gene

Incident ischemic stroke was defined as non-hemorrhagic stroke with a focal neurological deficit lasting ≥ 24 hours confirmed by the medical record or a focal or non-focal neurological deficit with positive imaging confirmed with medical records in participants without a prior history of stroke. Genomic DNA was analyzed using droplet digital PCR^6^ to determine *HBA* copy number. The covariates age, sex, race, health insurance (yes or no), highest education level obtained (less than high school, high school, some college, college or more), annual income (≤ $20K, $20-34K, $35-74K, ≥ $75K), and smoking status (categorized by never, past, or current smoker), history of hyperlipidemia, regular use of lipid lowering medication, and regular use of aspirin were self-reported. Self-reported use of lipid lowering medication was restricted to those also reporting hyperlipidemia. Region was defined as previously described in three geographic areas: stroke belt buckle, stroke belt, and stroke nonbelt.^11^ Atrial fibrillation was defined by self-report of a physician diagnosis or ECG evidence. The presence of left ventricular hypertrophy (LVH) was defined by 12-lead ECG. Hypertension was defined as systolic blood pressure >=130, diastolic blood pressure >=80, or self-reported current medication use to control blood pressure. Chronic kidney disease (CKD) was defined by the 2021 CKD-Epi creatinine-cysteine equation and estimated glomerular filtration rate less than 60mL/min/1.73m^2^, including those with end-stage kidney disease, and/or urine albumin to creatinine ratio ≥ 30 mg/g measured on urine collected during the baseline in-home examination. Fasting glucose levels ≥ 126 mg/dL, random glucose ≥ 200 mg/dL, or self-reported use of glucose-lowering medication was used to define diabetes mellitus. Hemoglobin, mean corpuscular volume (MCV), mean corpuscular hemoglobin (MCH), mean corpuscular hemoglobin concentration (MCHC), and red-cell distribution width-coefficient of variation (RDW-CV) values were measured or calculated from blood collected during the in-home examination. The first 10 principal components of ancestry were calculated from Infinium Expanded Multi-Ethnic Genotyping Array data on 7,032 (79%) participants. *HBA* regulatory single nucleotide polymorphisms (SNPs) (rs11248850, rs11865131, rs7203560)^12,13^ and a NOS3 SNP (rs1549758)^14^ were determined from these array data. The *HBA* regulatory SNPs rs11248850 and rs11865131 were in high linkage disequilibrium and were concordant in all but five participants. Therefore, we evaluated only rs11248850 and not rs11865131 in the models. The Framingham Stroke Risk Score (10-Year probability of stroke risk percentage)^15^ and Atherosclerosis Risk in Communities Study (ARIC) Stroke Risk Score, 10-year probability of ischemic stroke risk (percentage) were calculated among those who self-reported at baseline never having had a stroke.^16^

### Statistical methods

Multivariable Cox proportional hazards regression modeling was used to estimate the hazard ratio of *HBA* copy number on time to first ischemic stroke. Covariates included age, sex, region, insurance status, education level, income, hypertension, atrial fibrillation, left ventricular hypertrophy, smoking status, and diabetes mellitus. Pre-specified tests of interaction between *HBA* copy number and age, sex, and sickle cell trait were performed on fully adjusted models. Pre-specified sensitivity analyses were performed by adding each of the following to the model: hemoglobin, CKD, both hemoglobin and CKD, regular aspirin use, regular statin use, the first ten principal components of ancestry, and putative *HBA* regulatory single nucleotide polymorphisms rs11248850 and rs7203560. A post-hoc sensitivity analysis for the association of the rs1549758 NOS3 (nitric oxide synthase 3 or endothelial nitric oxide synthase) SNP with incident ischemic stroke was performed.

For the ischemic stroke time-to-event analysis, time to event was defined as the number of years between the initial in-home interview date and date D where D was the minimum of D1 and D2 which were defined as follows. D1 was the last follow-up date provided by REGARDS as the last time the participant was contacted for status. The outcome associated with this date was either “No Event” or “Death”. D2 was the date when the individual had an ischemic stroke. Individuals with a D2 date prior to their initial in-home interview were excluded from consideration in this paper. Individuals with a D2 date after interview but prior to D1 time were recorded with an ischemic stroke event and D = D2. Individuals with D2 after D1 had stroke onset after their last REGARDS follow-up and they were recorded with “No Event” and censored at date D = D1. This decision to censor stroke events occurring after the end of REGARDS follow-up avoids bias associated with having extended follow-up only for one type of subgroup - those having a stroke event.

Missing data for the primary outcome, secondary outcomes, and explanatory variables were typically rare (< 0.5%) with some exceptions, e.g., hemoglobin values (Table 1). Multiple imputation methods were used in the multivariable analyses. The R package “mice” Version 3.14.0 was used to create and analyze the resulting imputations.^17^

**Table 1.**
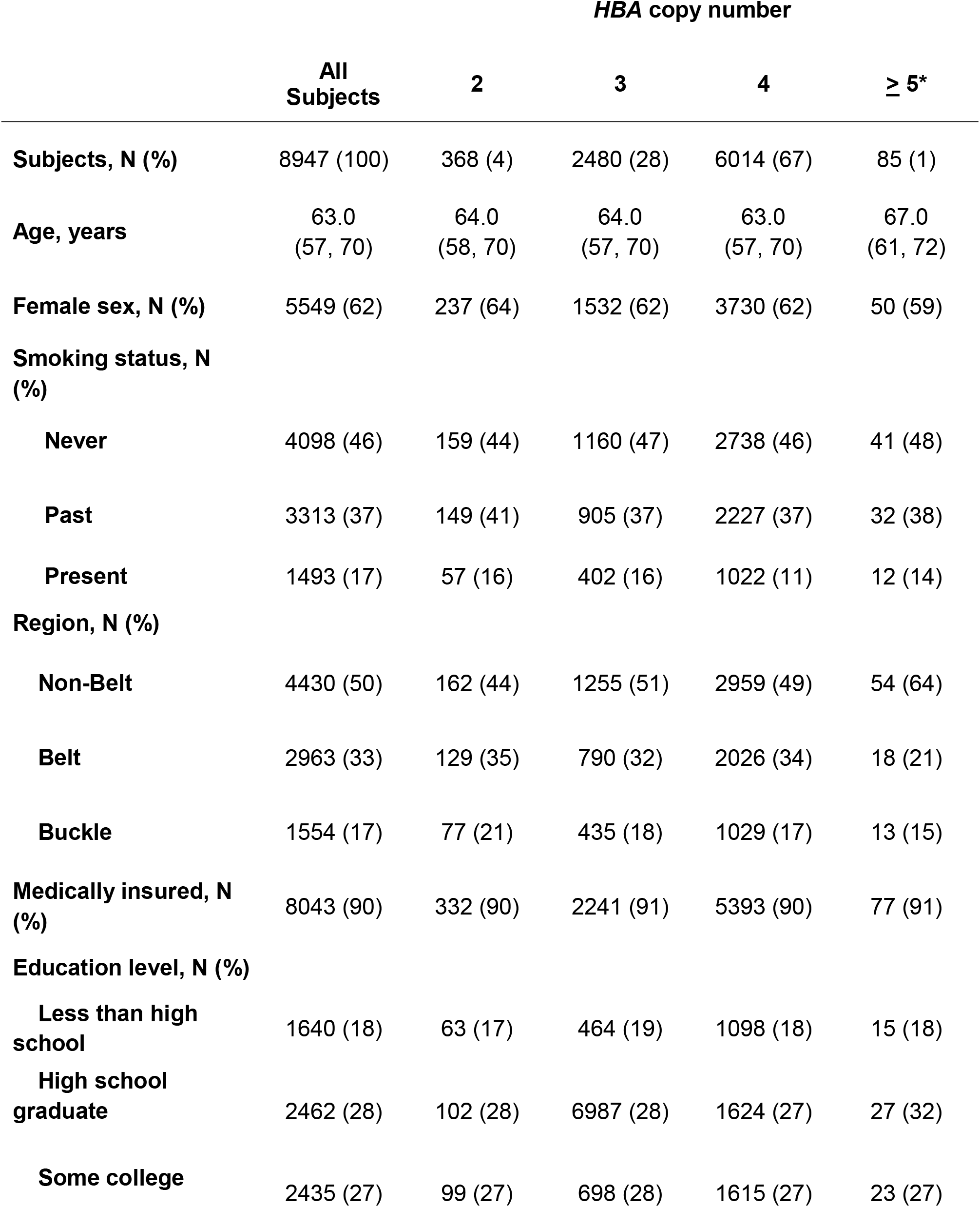
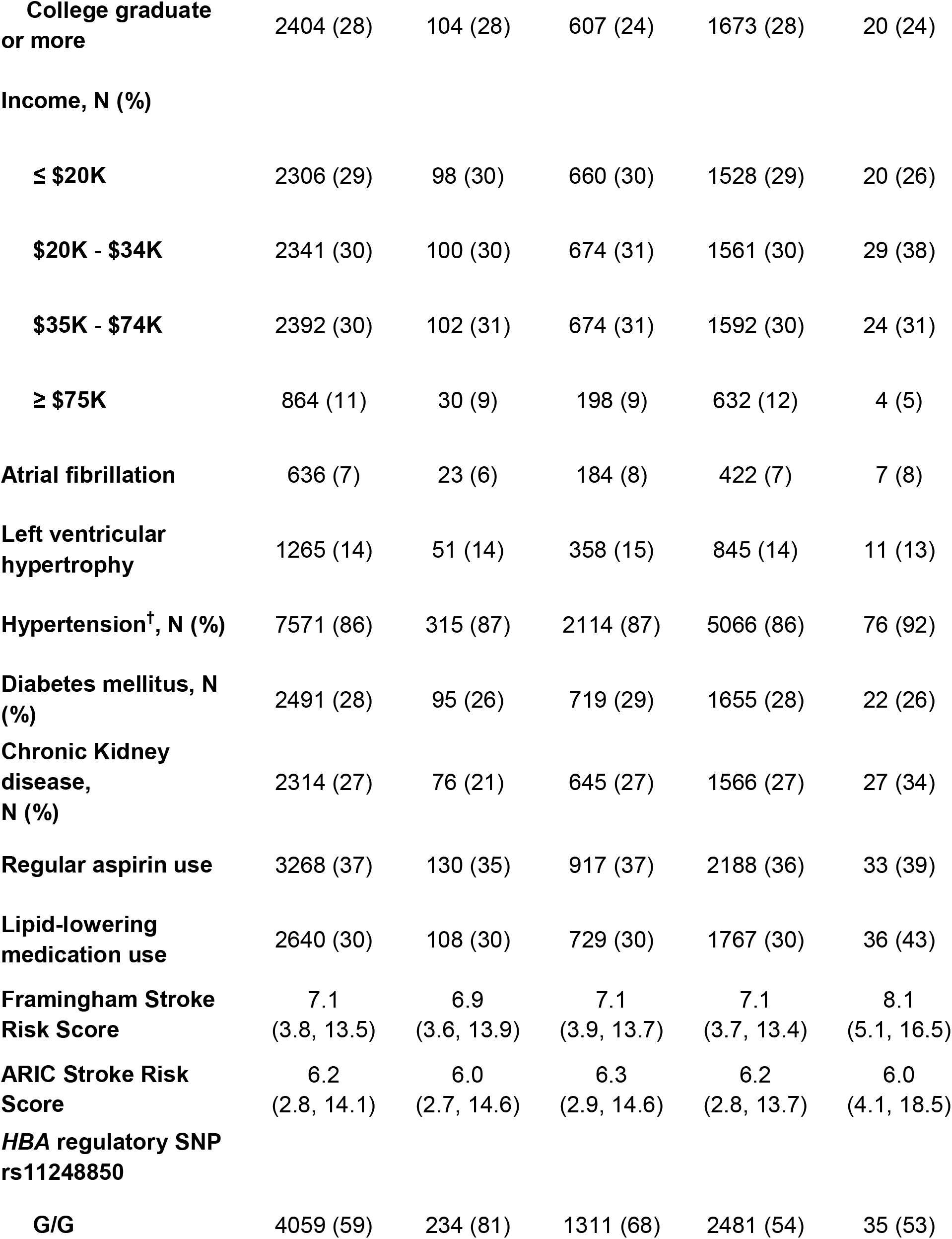
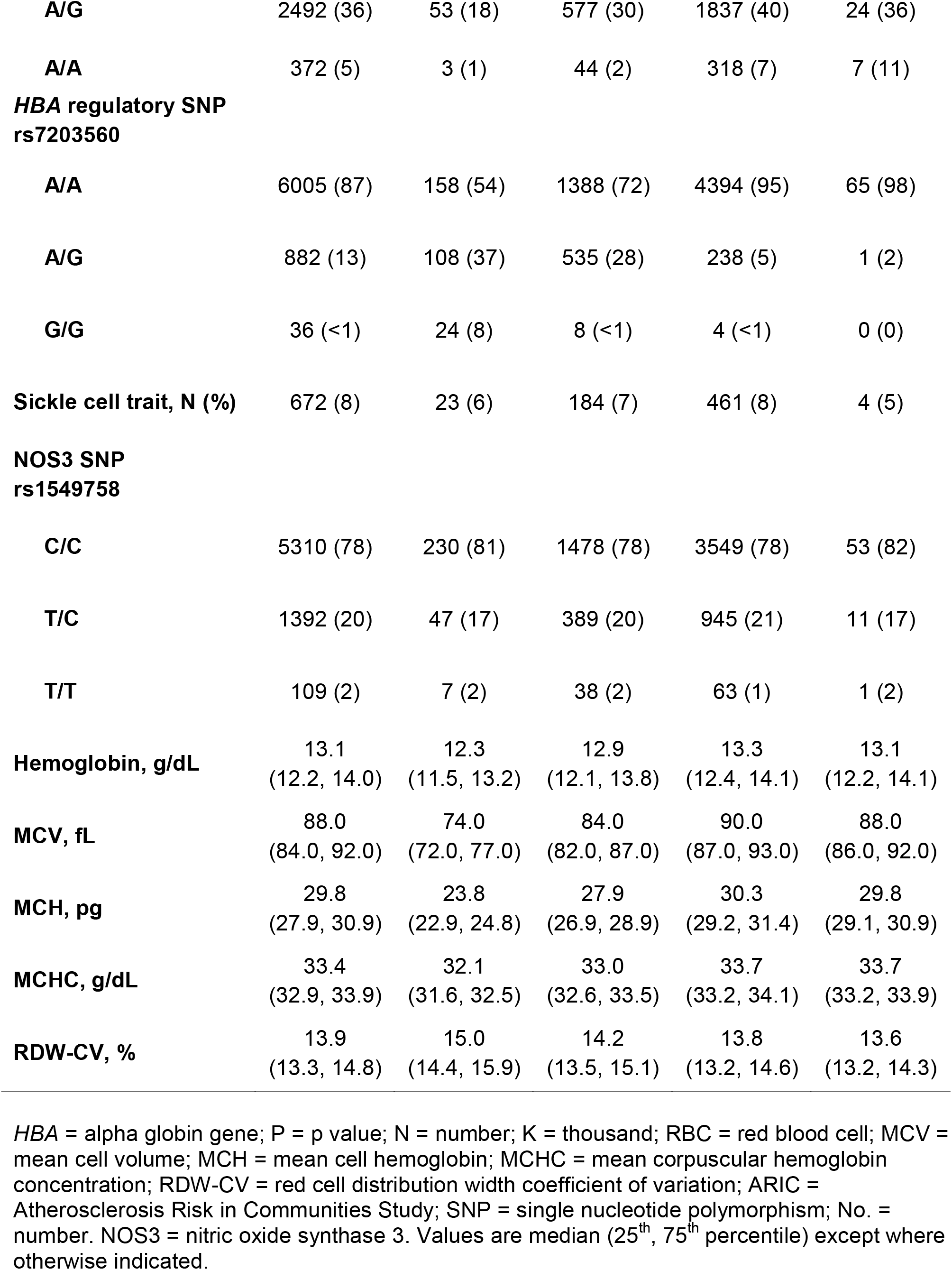
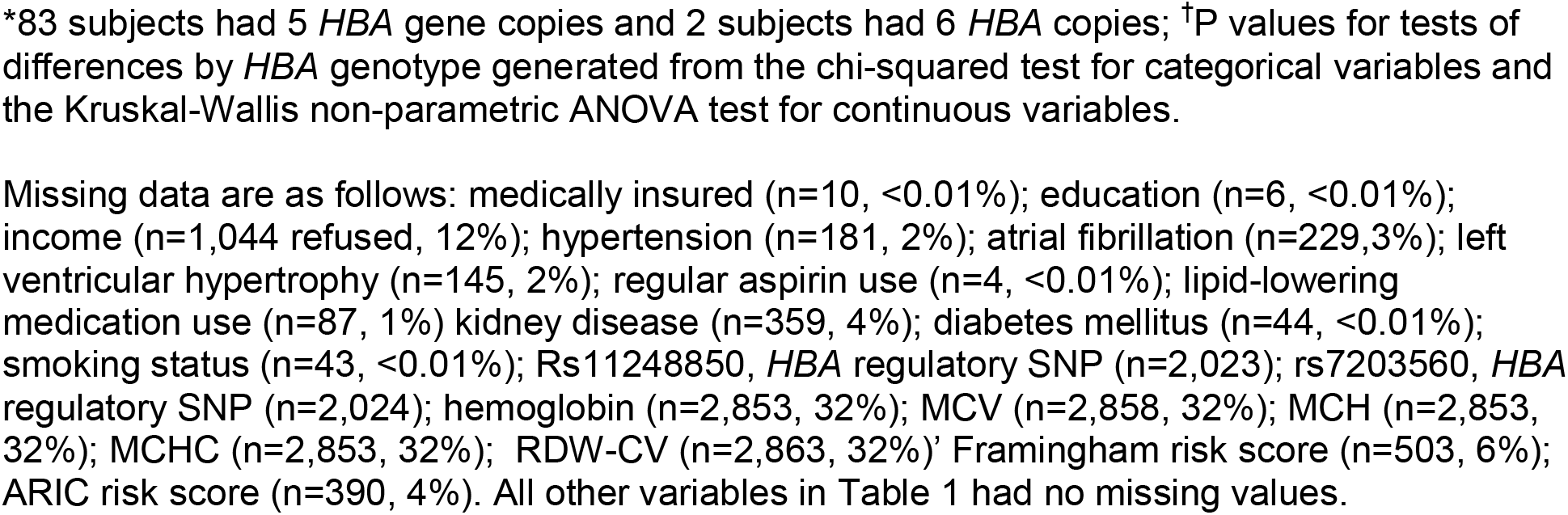
Clinical and demographic characteristics by *HBA* copy number

For diagnostic modeling of the model of ischemic stroke incidence using Cox proportional hazard techniques, Schoenfeld residuals were examined over follow-up time to detect violations of proportional hazards assumptions for the covariates in the analysis of ischemic outcomes. Examination of Schoenfeld residuals of the non-imputed and imputed data sets showed no suggestion of violation of proportional hazards for the regression covariates (p = 0.98).

For incident ischemic stroke, we performed an a priori power calculation based upon the Cox proportional-hazards model. We assumed an additive model where each additional *HBA* copy increased the hazard ratio for prevalent stroke by 25%. Given n=479 incident ischemic stroke events observed among those without baseline self-reported stroke, and the distribution of alpha globin alleles, the power to reject the hypothesis of no linear trend in *HBA* copy number on the log of the hazard rate is approximately 77%. In contrast, there were only 62 incident hemorrhagic stroke events and power to reject a linear trend in *HBA* copy number on the log of the hazard rate was only 13%; therefore, analysis of hemorrhagic stroke was not performed.

## RESULTS

Of the 8,947 REGARDs participants with available data (Figure 1), 479 (5.3%) participants had an incident ischemic stroke over a median (IQR) of 11.0 (5.7, 14.0) years’ follow-up. Of these participants with incident ischemic stroke, 393 (82%) were diagnosed with a non-hemorrhagic stroke with a focal neurological deficit lasting ≥ 24 hours and 86 (18%) were found to have a non-focal neurological deficit with positive imaging. There were 62 participants who developed incident hemorrhagic stroke. *HBA* gene copy number ranged from 2 to 6: 368 (4%) -α/-α, 2,480 (28%) -α/αα, 6,014 (67%) αα/αα, 83 (1%) ααα/αα and 2 (<1%) ααα/ααα (Table 1). The HR of ischemic stroke with *HBA* copy number, fully adjusted for age, sex, region, insurance status, education, income, hypertension, atrial fibrillation, LVH, smoking, and diabetes, was 1.04; 95%CI 0.89, 1.21; p = 0.66; (Table 2). There were no interactions between *HBA* copy number and age, sex, or sickle cell trait (Table 3). In pre-specified sensitivity analyses, the addition of hemoglobin, chronic kidney disease, aspirin use, statin use, or the first ten principal components of ancestry did not materially change the observed associations (Table 4). In a post-hoc sensitivity analysis we evaluated whether the putative NOS3 SNP was associated with incidence ischemic stroke in this population and no association was found (HR 1.03; 95%CI 0.84, 1.27; p = 0.79).

**Table 2.** Association of *HBA* copy number with incident ischemic stroke – fully adjusted analyses.

|  | **HR** | **95% CI** | **P value<sup>†</sup>** |
| --- | --- | --- | --- |
| ***HBA* copy number** | 1.04 | (0.89, 1.21) | 0.66 |
| **Age, per year** | 1.05 | (1.03, 1.06) | <0.001 |
| **Sex** |  |  |  |
| **Female (ref)<sup>§</sup>** |  |  |  |
| **Male** | 0.80 | (0.66, 0.96) | 0.02 |
| **Region** |  |  |  |
| **Non-Belt (ref)** |  |  |  |
| **Belt** | 1.15 | (0.94, 1.41) | 0.18 |
| **Buckle** | 1.19 | (0.92, 1.52) | 0.19 |
| **Medically insured** |  |  |  |
| **No (ref)** |  |  |  |
| **Yes** | 0.87 | (0.62, 1.21) | 0.41 |
| **Education level** |  |  |  |
| **< HS Grad (ref)** |  |  |  |
| **HS Grad** | 1.13 | (0.87, 1.45) | 0.36 |
| **Some College** | 0.78 | (0.59, 1.04) | 0.09 |
| **≥ College Grad** | 0.75 | (0.55, 1.02) | 0.07 |
| **Income** |  |  |  |
| **< \$20K (ref)** | | | |
| **\$20K - \$34K** | 1.09 | (0.85, 1.40) | 0.48 |
| **\$35K - \$74K** | 1.03 | (0.77, 1.37) | 0.83 |
| **≥ \$75K** | 0.72 | (0.45, 1.17) | 0.19 |
| **Hypertension** | 1.51 | (1.09, 2.08) | 0.01 |
| **Atrial fibrillation** | 1.48 | (1.09, 1.99) | 0.01 |
| **Left ventricular hypertrophy** | 1.32 | (1.04, 1.67) | 0.02 |
| **Smoking status** |  |  |  |
| **Never (ref)** |  |  |  |
| **Past** | 1.04 | (0.84, 1.26) | 0.79 |
| **Present** | 1.62 | (1.25, 2.10) | <0.001 |
| **Diabetes mellitus** |  |  |  |
| **No (ref)** |  |  |  |
| **Yes** | 1.67 | (1.39, 2.01) | <0.001 |

*HBA* = alpha globin gene; HR = hazard ratio; CI = confidence interval; ref = reference; HS = high school. Grad = graduate; K = thousand.
<sup>†</sup>P values were generated with a Cox proportional hazards multivariable regression employing a linear effect (i.e., additive model for risk) of *HBA* allele count on the log of the hazard ratio. All variables in the table were included in the multivariable model. Multiple imputations were performed for missing data.

**Table 3.**
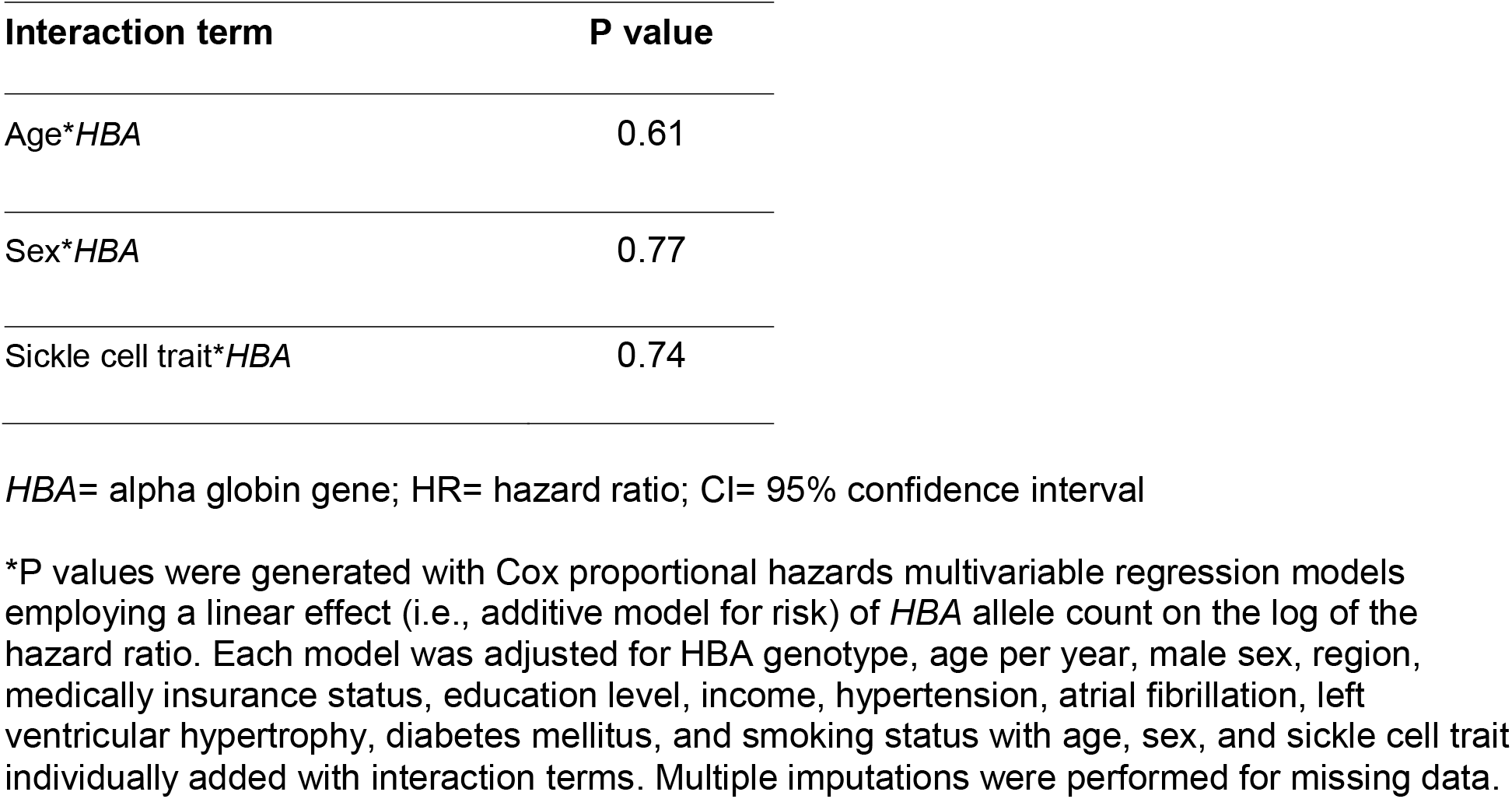
Pre-specified tests for interaction between *HBA* genotype and age, sex, and sickle cell trait on incident ischemic stroke in fully adjusted models

**Table 4.**
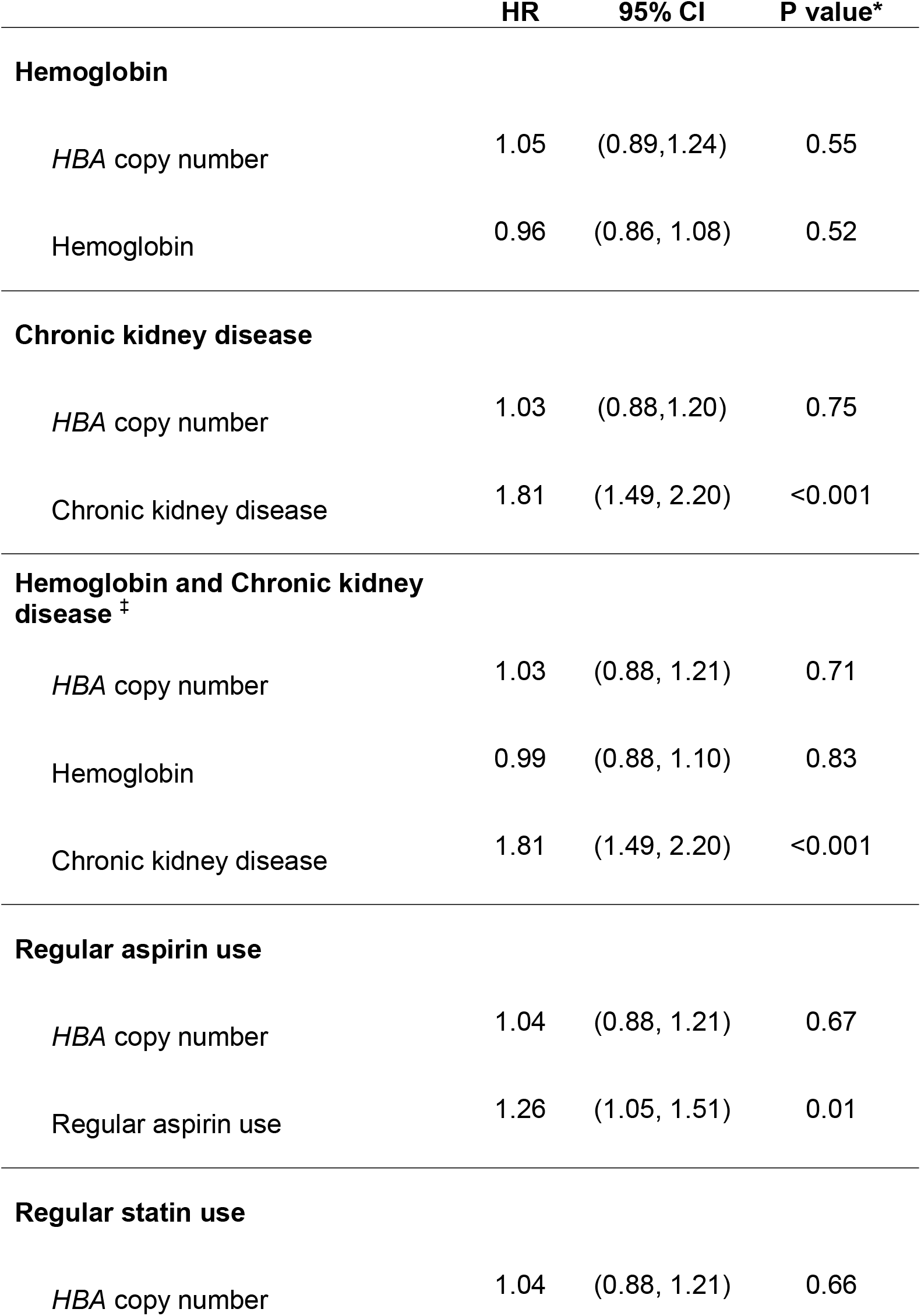
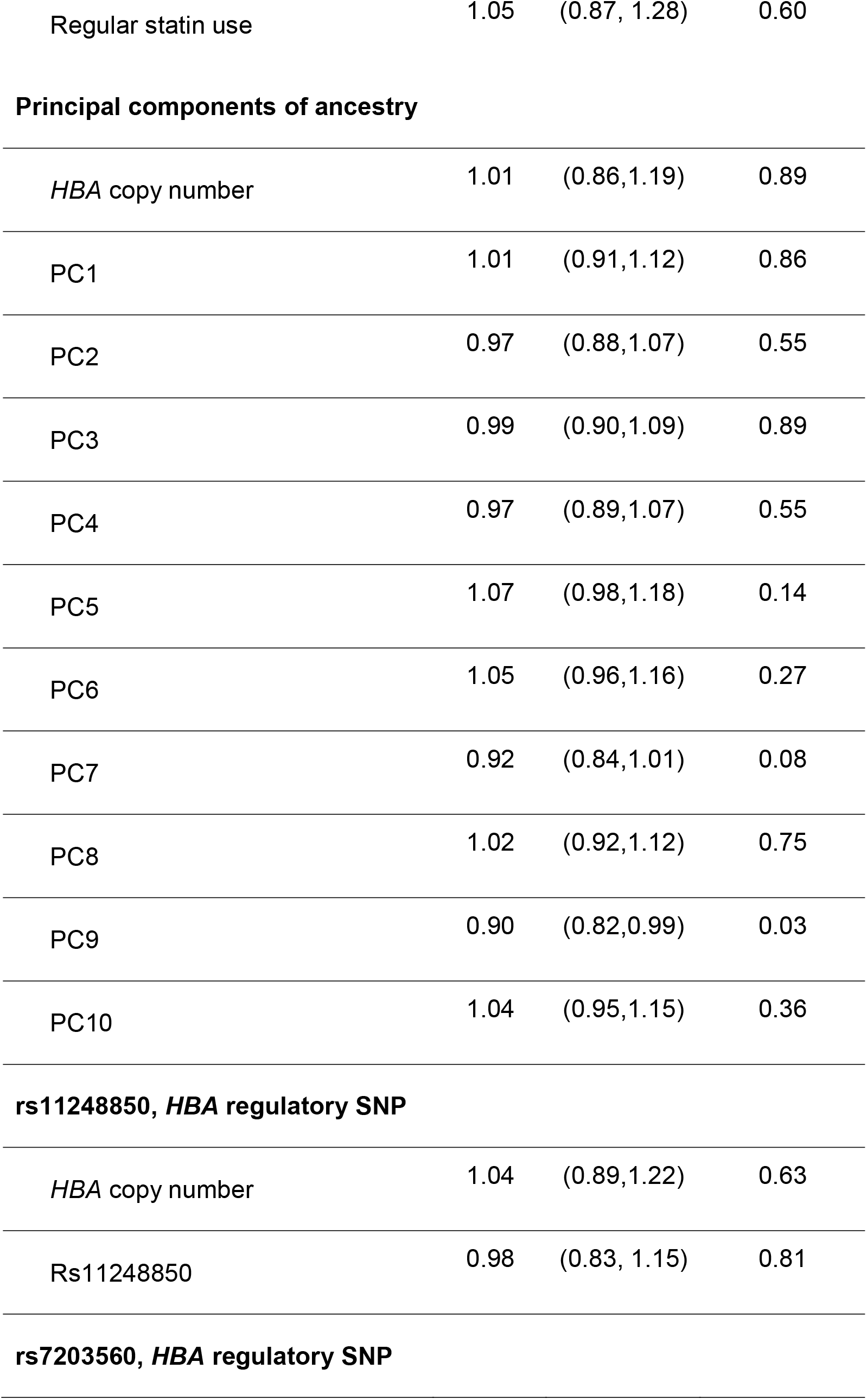
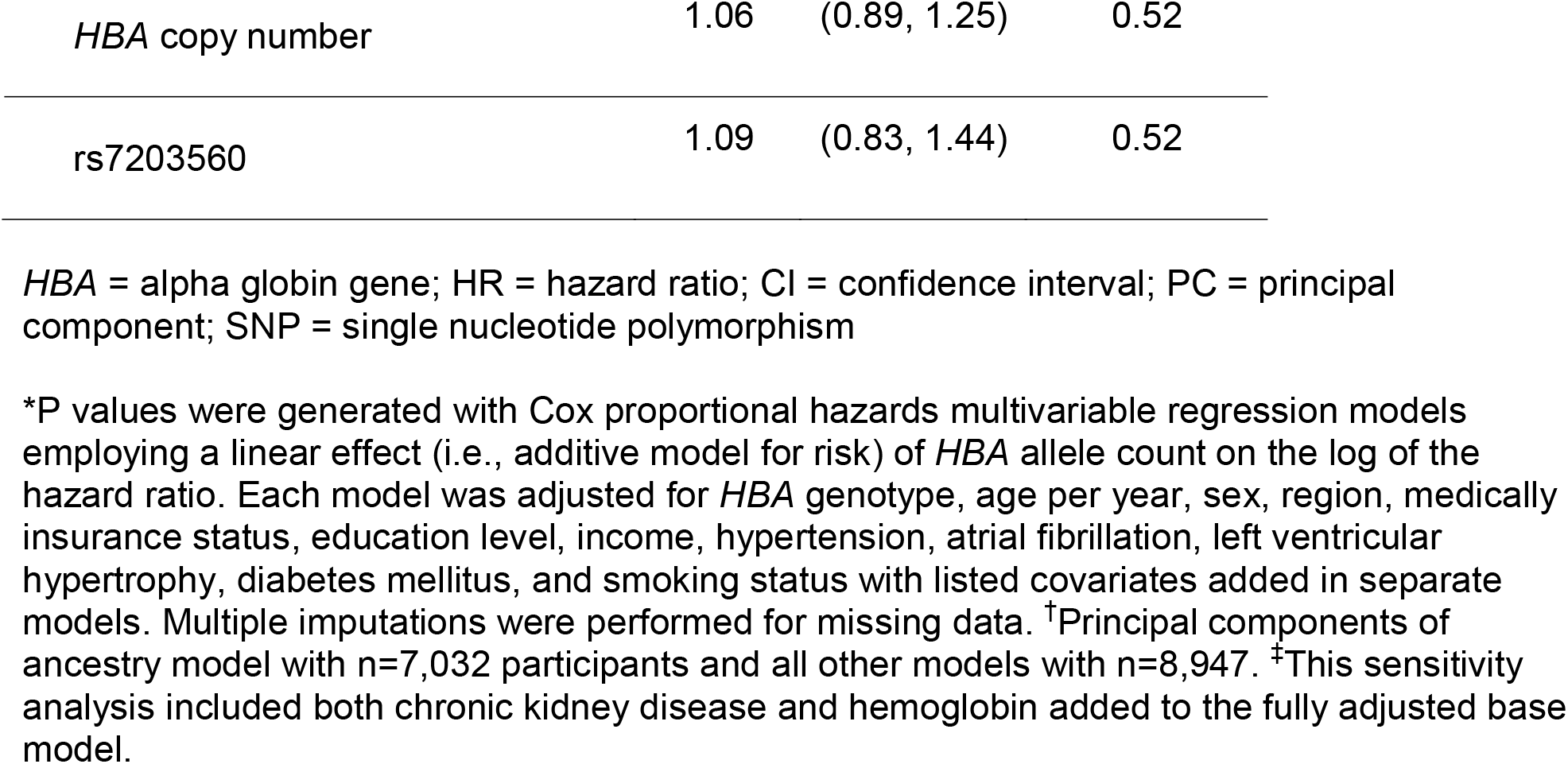
Pre-specified sensitivity analyses for the association of *HBA* genotype with incident ischemic stroke – fully adjusted models with the separate addition of covariates for each model.

|  | HR | 95% CI | P value* |
| --- | --- | --- | --- |
| **Hemoglobin** |  |  |  |
| *HBA* copy number | 1.05 | (0.89,1.24) | 0.55 |
| Hemoglobin | 0.96 | (0.86, 1.08) | 0.52 |
| **Chronic kidney disease** |  |  |  |
| *HBA* copy number | 1.03 | (0.88,1.20) | 0.75 |
| Chronic kidney disease | 1.81 | (1.49, 2.20) | <0.001 |
| **Hemoglobin and Chronic kidney disease<sup>‡</sup>** |  |  |  |
| *HBA* copy number | 1.03 | (0.88, 1.21) | 0.71 |
| Hemoglobin | 0.99 | (0.88, 1.10) | 0.83 |
| Chronic kidney disease | 1.81 | (1.49, 2.20) | <0.001 |
| **Regular aspirin use** |  |  |  |
| *HBA* copy number | 1.04 | (0.88, 1.21) | 0.67 |
| Regular aspirin use | 1.26 | (1.05, 1.51) | 0.01 |
| **Regular statin use** |  |  |  |
| *HBA* copy number | 1.04 | (0.88, 1.21) | 0.66 |
| Regular statin use | 1.05 | (0.87, 1.28) | 0.60 |
| Principal components of ancestry |  |  |  |
| *HBA* copy number | 1.01 | (0.86,1.19) | 0.89 |
| PC1 | 1.01 | (0.91,1.12) | 0.86 |
| PC2 | 0.97 | (0.88,1.07) | 0.55 |
| PC3 | 0.99 | (0.90,1.09) | 0.89 |
| PC4 | 0.97 | (0.89,1.07) | 0.55 |
| PC5 | 1.07 | (0.98,1.18) | 0.14 |
| PC6 | 1.05 | (0.96,1.16) | 0.27 |
| PC7 | 0.92 | (0.84,1.01) | 0.08 |
| PC8 | 1.02 | (0.92,1.12) | 0.75 |
| PC9 | 0.90 | (0.82,0.99) | 0.03 |
| PC10 | 1.04 | (0.95,1.15) | 0.36 |
| rs11248850, *HBA* regulatory SNP |  |  |  |
| *HBA* copy number | 1.04 | (0.89,1.22) | 0.63 |
| Rs11248850 | 0.98 | (0.83, 1.15) | 0.81 |
| rs7203560, *HBA* regulatory SNP |  |  |  |
| *HBA* copy number | 1.06 | (0.89, 1.25) | 0.52 |
| rs7203560 | 1.09 | (0.83, 1.44) | 0.52 |

*HBA* = alpha globin gene; HR = hazard ratio; CI = confidence interval; PC = principal component; SNP = single nucleotide polymorphism
\*P values were generated with Cox proportional hazards multivariable regression models employing a linear effect (i.e., additive model for risk) of *HBA* allele count on the log of the hazard ratio. Each model was adjusted for *HBA* genotype, age per year, sex, region, medically insurance status, education level, income, hypertension, atrial fibrillation, left ventricular hypertrophy, diabetes mellitus, and smoking status with listed covariates added in separate models. Multiple imputations were performed for missing data. <sup>†</sup>Principal components of ancestry model with n=7,032 participants and all other models with n=8,947. <sup>‡</sup>This sensitivity analysis included both chronic kidney disease and hemoglobin added to the fully adjusted base model.

## DISCUSSION

Although a reduction in *HBA* copy number is expected to increase endothelial nitric oxide signaling in the human vascular endothelium, *HBA* copy number was not associated with incident ischemic stroke in this large longitudinal cohort of Black Americans. This finding is consistent with a prior study demonstrating no association between *HBA* copy number and hypertension but differs from a study demonstrating an association with chronic and end-stage kidney disease in which *HBA* gene deletions are protective.^6,18^ Together, these population studies suggest that the physiological roles of alpha globin may differ between the renal and cerebral vascular beds.

The strengths of this study include a robust measurement of *HBA* copy number with ddPCR with the analysis performed in a well-characterized, large, national cohort of Black Americans with clearly defined cerebrovascular outcomes followed for an extensive period. Moreover, we adjusted for key stroke risk factors including demographic, social, and biomedical variables to address the novel question of whether *HBA* copy number is associated with stroke risk. We explored the relevance of the NOS3 SNP rs1549758 and found no association between this SNP and incident ischemic stroke in a fully adjusted model. This replicates the finding of no association with stroke among participants with African ancestry in a prior GWAS study.^14^ Given the absence of a significant main effect for either *HBA* CNV or the NOS3 SNP on incident ischemic stroke in this cohort, we did not pursue a model with both main effects nor did we test for interaction.

Our study has limitations, including that the population is limited to Black Americans and the associations of *HBA* copy number and ischemic stroke risk may vary in other populations with *HBA* copy number variation and outside of the U.S. We did not have sufficient power to evaluate other stroke subtypes such as hemorrhagic stroke, though increases in vascular NO signaling could affect bleeding risk through its effects on platelets. An evaluation of the association of *HBA* copy number and hemorrhagic stroke risk could be evaluated in other populations with a higher frequency of *HBA* copy number variation. For example, in Sub-Saharan Africa, there is a slightly higher frequency of hemorrhagic stroke than reported than in the U.S.^19^

## CONCLUSION

In a large longitudinal cohort of Black American adults, we found that *HBA* copy number was not associated with incident ischemic stroke.

## Data Availability

Data are available from the University of Alabama.

## DATA AVAILABILITY

Data are available from the University of Alabama at Birmingham. The genetic data is available in dbGaP with study accession number phs002719.v1.p1.

